# Time until symptoms and design-related associations in Alzheimer’s disease clinical progression analyses

**DOI:** 10.64898/2026.05.10.26352825

**Authors:** Kellen K. Petersen, Yan Li, Suzanne E. Schindler

## Abstract

**Introduction:** Studies of the risk and timing of symptomatic Alzheimer’s disease (AD) in cognitively unimpaired individuals are challenging due to the relatively small number of clinical progressors and limited clinical follow-up, which can lead to design-related associations. Clock models can be used to anchor the timing of events to biological events such as biomarker positivity. We hypothesized that estimated age at plasma %p-tau217 positivity based on clock models is less affected by design-related associations as compared to baseline age.

**Methods:** Data from the Knight Alzheimer Disease Research Center (Knight ADRC) and Alzheimer’s Disease Neuroimaging Initiative (ADNI) were analyzed. Age at %p-tau217 positivity was estimated using two clock model approaches, TIRA and SILA. The C-index of estimated age at plasma %p-tau217 positivity and age at the baseline plasma sample (baseline age) for ranking age of AD symptom onset was evaluated in initially cognitively unimpaired individuals, including progressors and non-progressors. In progressor sub-cohorts, baseline age and time from %p-tau217 positivity to baseline were associated with time from baseline until symptom onset; baseline age and estimated age at %p-tau217 positivity were associated with age at symptom onset. Commonality analyses partitioned the variance unique to each predictor and shared between predictors. Randomization analyses evaluated whether observed associations exceeded those expected by chance.

**Results:** Estimated age at %p-tau217 positivity enabled analyses of a greater number of progressors in the research cohorts, which did not have plasma %p-tau217 data from every clinical assessment. The estimated age at %p-tau217 positivity had a higher C-index than baseline age for ordering the likelihood of AD symptom onset when all follow-up was considered; when follow-up was truncated, the C-index for estimated age at %p-tau217 positivity remained stable while the C-index for baseline age became inflated. In progressors, estimated age at %p-tau217 positivity contributed unique variance beyond baseline age in associations with age at symptom onset. Randomization analyses in the larger Knight ADRC found that associations between clock-derived measures and time from baseline until symptom onset and age at symptom onset exceeded the permuted null distribution, with some mixed results in the smaller ADNI cohort.

**Conclusions:** Compared to baseline age, the biologically-anchored estimated age at %p-tau217 positivity is less susceptible to design-related associations and incrementally improves prediction of age at symptom onset in analyses conditional on progression.

## Introduction

A key goal of Alzheimer’s disease (AD) research is to identify cognitively unimpaired individuals who are at high risk for developing symptomatic AD with the goal of intervening to slow or prevent the onset of AD symptoms ^1-3^. A major challenge for AD clinical trials is enrolling cognitively unimpaired participants who are likely to develop symptoms within a relatively short follow-up period. Notably, clinical trials in families with autosomal dominant AD mutations are more efficient due to enrollment of individuals with a relatively predictable age at AD symptom onset based on their family history ^3-5^. Clinical trials in sporadic AD may benefit from biomarkers and methodological approaches that predict both the risk and timing of AD symptoms.

Studies of AD clinical progression are especially challenging due to the relatively small number of clinical progressors and limited clinical follow-up, requiring large cohorts for adequate statistical power. Clock models enable AD-related events and trajectories to be anchored to a distinct biological event such as amyloid positivity, enabling a timeline to be constructed that can be projected to even before the period of clinical observation ^6^. Recently, Petersen *et al*. used plasma %p-tau217 clock models to estimate the age at %p-tau217 positivity ^7^. However, because models of the absolute timing of symptomatic AD were developed only in the progressor sub-cohorts and onset of symptomatic AD must occur after %p-tau217 positivity, design-related associations may occur as described by Insel and Donohue ^8^.

In this study we examined whether associations of estimated %p-tau217 positivity with time from baseline until onset of symptomatic AD and age at symptom onset are less affected by design-related associations as compared to associations with baseline age. To compare these predictors and outcomes, we used the C-index, regression models, commonality analyses, and permutation-based randomization analyses. Further, changes in associations with follow-up truncation were examined.

## Methods

### Participants

All protocols were approved by the Washington University in St. Louis Institutional Review Board (Human Research Protection Office) and by the local Institutional Review Boards at each participating ADNI site. Written informed consent was obtained from every participant or, when appropriate, from a legally authorized representative.

Data from Petersen *et al*. were analyzed and included participants from two cohorts: the Knight Alzheimer Disease Research Center (Knight ADRC) and the Alzheimer’s Disease Neuroimaging Initiative (ADNI) ^7^. The age at %p-tau217 positivity was estimated using two methods, TIRA and SILA ^7^. C-index analyses included initially cognitively unimpaired individuals with both clinical progressors and non-progressors that have previously been described ^7^. Regression, commonality, and randomization analyses included only clinical progressors to symptomatic AD as defined by both the clinical syndrome and AD biomarkers: individuals who were initially cognitively unimpaired and developed cognitive impairment with an AD syndrome after estimated plasma %p-tau217 positivity, and who had an AD syndrome at their last assessment.

### Variables

Estimated age at %p-tau217 positivity was derived from the TIRA or SILA clock models as previously described ^7^. The age at the first/baseline plasma sample included in the study was considered baseline age. The age at symptom onset was the age at first diagnosis of cognitive impairment due to AD. Two outcomes were examined: 1) time from baseline until symptom onset (age at symptom onset minus baseline age), and 2) age at symptom onset.

### C-index analyses

For C-index analyses, the outcomes were defined as age at either progression to an AD syndrome regardless of %p-tau217 positivity status or age at an AD syndrome following estimated %p-tau217 positivity (symptomatic AD). The predictors were either baseline age or estimated age at %p-tau217 positivity. The C-index was computed as a concordance measure in which a pair of individuals was considered comparable only when their event intervals were strictly non-overlapping, ensuring that temporal ordering could be established unambiguously. Bootstrapped 95% confidence intervals for the C-index were obtained using 2,000 bootstrap samples with replacement. As a sensitivity analysis, interval-censored event times for progressors were replaced with interval midpoints and C-indices were re-estimated using standard right-censored Cox models with 95% confidence intervals derived from the analytical variance of the concordance statistic.

To simulate fixed-duration observation windows, follow-up was truncated at fixed intervals from the baseline plasma sample. Only progression that was observed before the end of the truncation window was considered as an event. Follow-up times of ≤17, ≤14, ≤11, ≤8, ≤5, and ≤2 years from the baseline plasma sample were evaluated in addition to the full available follow-up.

### Regression analyses

In the subset of individuals who progressed to symptomatic AD, two outcomes were examined: time from baseline until symptom onset (age at symptom onset minus baseline age), and age at symptom onset. For each outcome, three linear regression models were fit using baseline age alone, a second predictor alone, and both predictors combined, with adjusted R^2^ reported for each. For age at symptom onset, the second predictor was estimated age at %p-tau217 positivity. For time from baseline until symptom onset, the second predictor was time from estimated %p-tau217 positivity to baseline, which anchored the clock derived measure to baseline age. The incremental contribution of each predictor was evaluated using F-tests. The non-nested comparison between baseline age alone and the second predictor alone used the Williams test for dependent, overlapping correlations, implemented using the cocor package in R.

To examine the sensitivity of associations to length of follow-up, progressors were progressively excluded based on time from baseline to symptom onset. Follow-up was restricted to ≤17, ≤14, ≤11, ≤8, ≤5, and ≤2 years, retaining only progressors whose symptom onset occurred within each threshold. At each restriction level, simple linear regressions of each outcome on each predictor were fit and adjusted R^2^ was recorded. This was performed separately for each of the four cohort/method progressor datasets.

### Commonality analyses

Commonality analyses were used to partition the total R^2^ of a two-predictor regression model into components uniquely attributable to each predictor and the component common to both. In a two-predictor model, the unique contribution of one predictor is defined as the difference in R^2^ between the full model and the model containing the other predictor. The common component is the remainder of total R^2^ after subtracting the R^2^ from both unique contributions and represents variance that cannot be attributed to either predictor independently. Unlike standardized regression coefficients, commonality analysis provides an additive decomposition of the total R^2^. For each outcome and cohort/method combination, commonality coefficients were computed using the yhat package in R.

### Randomization analysis

To evaluate whether observed associations between each predictor and each outcome exceeded those obtained by a structurally similar but uninformative predictor, permutation-based randomization analyses were performed for two outcomes: time from baseline until symptom onset and age at symptom onset. Predictor values were randomly shuffled across individuals, preserving the observed distribution of values while disrupting any true predictor-outcome relationship. Shuffling procedures were tailored to each predictor-outcome combination to preserve relevant mathematical relationships under the null.

For time from baseline until symptom onset, the predictor was time from %p-tau217 positivity to baseline. Estimated age at %p-tau217 positivity was shuffled across participants, and time from %p-tau217 positivity to baseline was recomputed as baseline age minus the shuffled value, with the outcome held fixed.

For age at symptom onset, three predictors were evaluated. When the predictor was estimated age at %p-tau217 positivity, two null approaches were used. In the first, estimated age at %p-tau217 positivity was shuffled directly across participants with the outcome held fixed, providing a less constrained null that does not preserve the baseline age structure of the predictor. In the second, the time from %p-tau217 positivity to baseline was shuffled across participants and the predictor was recomputed as observed baseline age minus the shuffled timing value, with the outcome held fixed, preserving the structural constraint that the null predictor remains anchored to baseline age. When the predictor was baseline age, values were shuffled directly across participants with the outcome unchanged.

In all cases, adjusted R^2^ was recomputed from a simple linear regression of the outcome on the shuffled predictor across 10,000 permutations to generate a null distribution. An empirical two-sided p-value was computed as the proportion of permuted adjusted R^2^ values deviating from the null mean by at least as much as the observed value, with the observed statistic included in the reference distribution ^9^. For most analyses the null distribution is centered near zero; for one analysis in which the shuffle procedure induces a structural relationship between predictor and outcome, centering ensures the p-value reflects deviation from the structural null rather than from zero ^9^. Analyses were performed separately in each dataset.

### Data and code availability

Data from the Knight ADRC can be requested by qualified investigators (https://knightadrc.wustl.edu/professionals-clinicians/request-center-resources/). Data from ADNI can be requested via the LONI website (https://adni.loni.usc.edu/). All analyses were performed in R (version 4.4.1). Codes used in these analyses are available from https://github.com/WashUFluidBiomarkers/design-related-associations-in-AD-clock-models.

## Results

### Cohort characteristics

The characteristics of the cohorts, including clinical progressors and non-progressors used to assess associations with risk for progression, have been previously described ^7^. The number of individuals with %p-tau217 values within the range of values that estimated time from %p-tau217 positivity could be estimated varied slightly by clock model: Knight ADRC, n=59 for TIRA and n=61 for SILA; ADNI, n=20 for TIRA and n=22 for SILA (**Figure 1**). Notably, not all clinical visits had plasma samples with measurements of %p-tau217 and some individuals who were cognitively unimpaired at their baseline clinical assessment developed symptoms before their first/baseline plasma sample (referred to as baseline age): Knight ADRC/TIRA 8 of 59 (13.6%), Knight ADRC/SILA 7 of 61 (11.5%), ADNI/TIRA 3 of 20 (15.0%), ADNI/SILA 3 of 22 (13.6%). While these additional individuals identified as progressors using estimated age at %p-tau217 positivity would not have been included as progressors in traditional analyses anchored on the baseline plasma sample, they were included in all analyses in the current study to provide a matched comparison.

**Figure 1.**
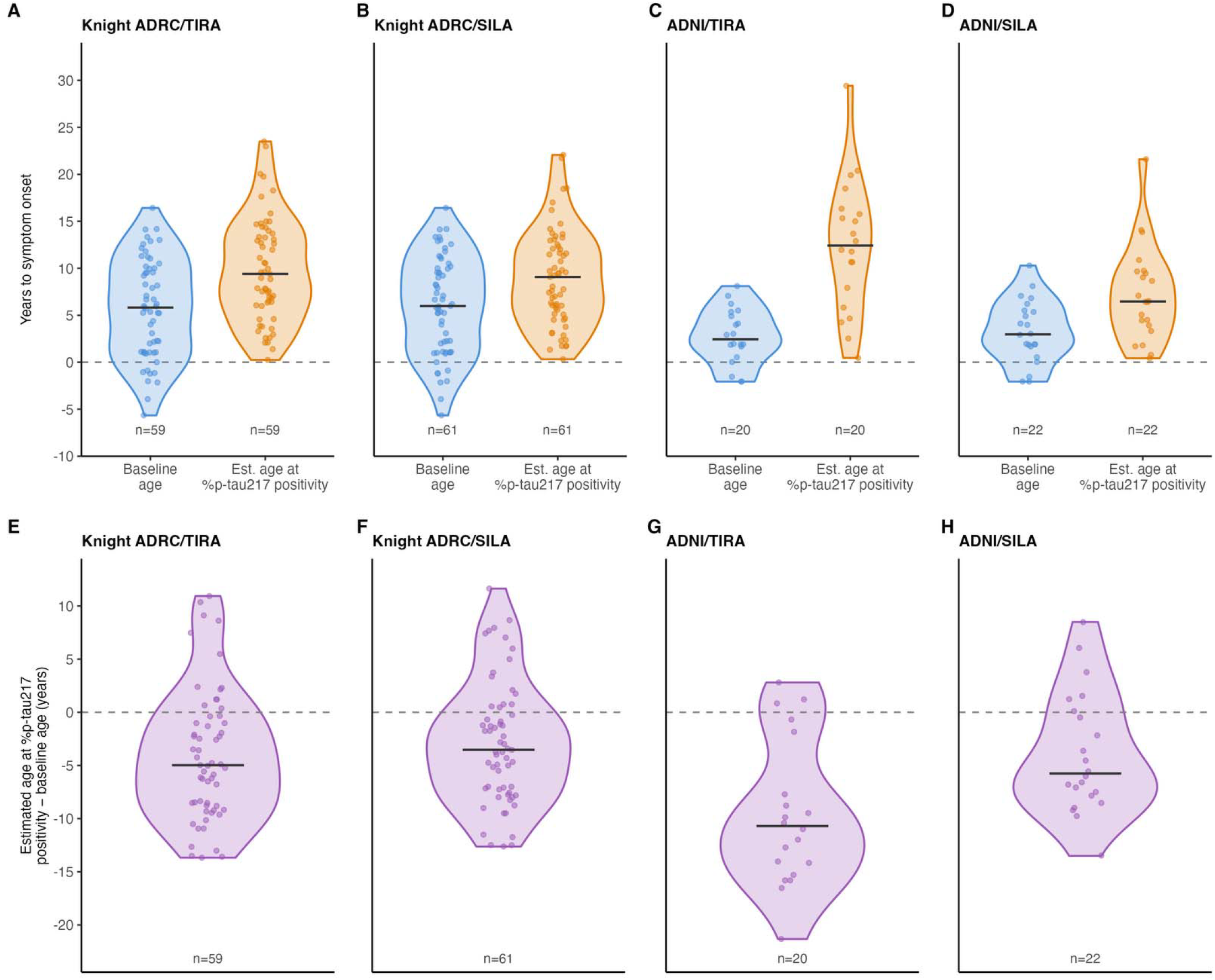
Distribution of years from baseline age or estimated age at %p-tau217 positivity to symptom onset in progressors. Distributions of years from baseline age (blue) or estimated age at %p-tau217 positivity (orange) **(A-D)** to symptom onset and estimated age at %p-tau217 positivity minus baseline age (purple) **(E-H)** for different cohort/method combinations. For **E-H**, negative values indicate that estimated %p-tau217 positivity occurred prior to the baseline plasma collection. Solid horizontal lines indicate medians.

The time until symptom onset in progressors was significantly longer when estimated age at %p-tau217 positivity was used as the anchor instead of baseline age (paired Wilcoxon signed-rank test, all p≤0.004; **Supplementary Table 1**). The median time from estimated age at %p-tau217 positivity to age at symptomatic AD was the following: Knight ADRC/TIRA 9.4 years, Knight ADRC/SILA 9.1 years, ADNI/TIRA 12.4 years, ADNI/SILA 6.5 years. In contrast, the median time from baseline age until age at symptom onset was the following: Knight ADRC/TIRA 5.8 years, Knight ADRC/SILA 6.0 years, ADNI/TIRA 2.4 years, ADNI/SILA 3.0 years.

The median baseline age was approximately 73 years and similar across cohorts/methods ^7^. The median clinical follow-up was slightly longer in the Knight ADRC (approximately 13 years) than ADNI (approximately 10 years) ^7^. Across all cohorts and methods, the sample was approximately 50% female ^7^.

### Concordance of age at symptom onset with age at %p-tau217 positivity or baseline age with follow-up truncation

In analyses of all individuals who were cognitively unimpaired at their baseline cognitive assessment, including both progressors and non-progressors, the ability of estimated age at %p-tau217 positivity or baseline age to rank age of onset for an AD syndrome or symptomatic AD (an AD syndrome with estimated %p-tau217 positivity) was evaluated using a C-index. When all available clinical follow-up was considered, the estimated age at %p-tau217 positivity demonstrated a higher concordance with age at onset of either an AD syndrome or symptomatic AD compared to baseline age (**Figure 2, Supplementary Figures 1–3**). As clinical follow-up was progressively truncated, the C-index for estimated age at %p-tau217 positivity remained stable across all observation windows. In contrast, the C-index for baseline age increased with progressive follow-up truncation at observation windows of ≤5 and ≤2 years. Consistent findings were obtained in the Knight ADRC cohort for both TIRA (**Figure 2**) and SILA (**Supplementary Figure 1**). In ADNI, the C-index for baseline age had greater inflation and was similar to estimated age at %p-tau217 positivity at the shortest observation windows for both TIRA (**Supplementary Figure 2**) and SILA (**Supplementary Figure 3**). In a sensitivity analysis using interval midpoints as the imputed event time and standard right-censored models, results were consistent with the primary interval-censored analysis (**Supplementary Figure 4**).

**Figure 2.**
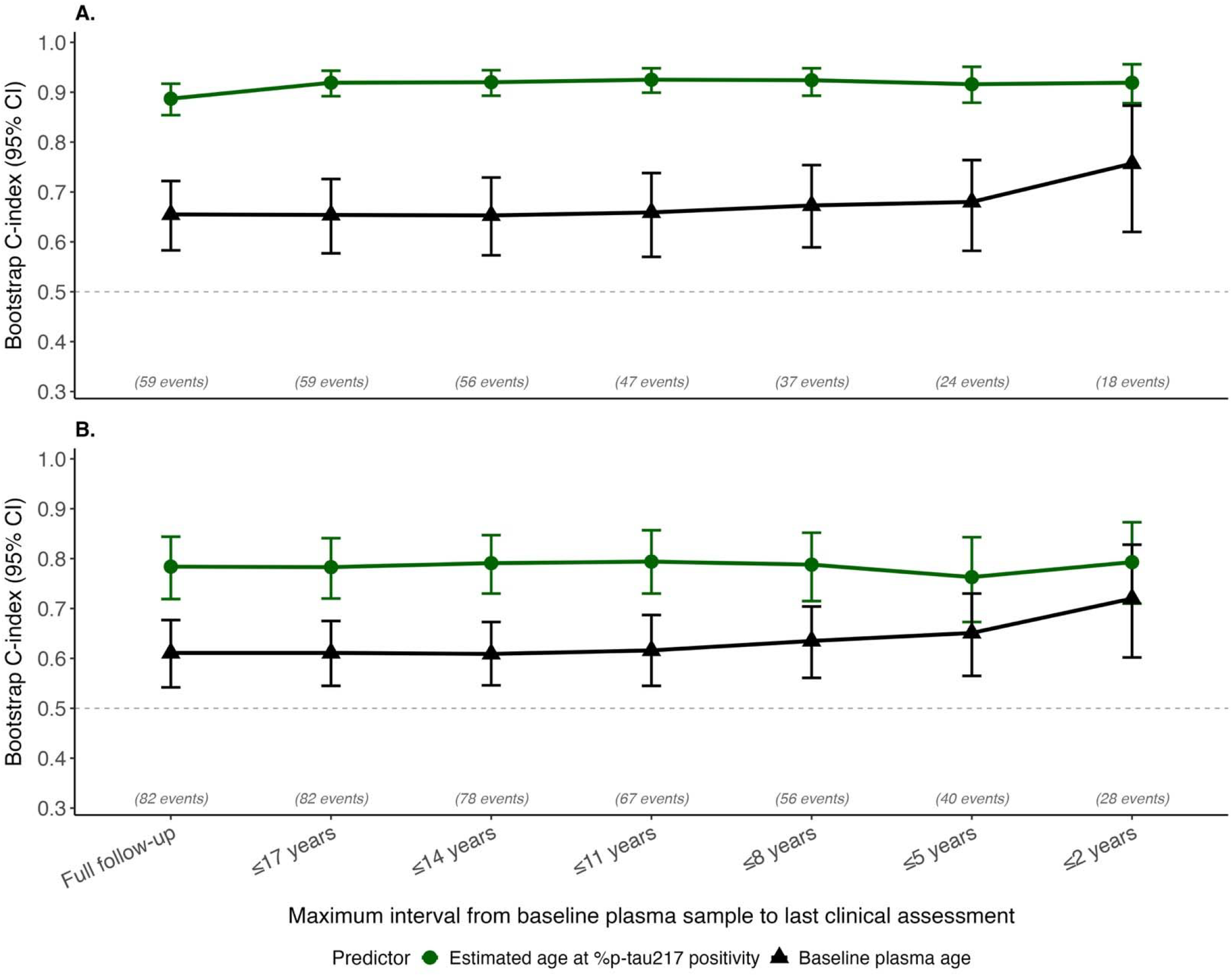
Concordance of age at symptom onset with age at %p-tau217 positivity or baseline age (Knight ADRC/TIRA). For individuals who were cognitively unimpaired at their baseline cognitive assessment, including both progressors and non-progressors, the ability of baseline age and estimated age at %p-tau217 positivity to rank age of onset of either **(A)** symptomatic AD (an AD syndrome with estimated %p-tau217 positivity) or **(B)** an AD syndrome were evaluated. Follow-up was truncated at fixed intervals from the baseline plasma sample to simulate trial observation windows of varying duration. The C-index was estimated using interval-censoring with 2000-sample bootstrapped 95% confidence intervals. Sample sizes and event counts at each restriction level are shown below the x-axis.

### Associations of outcomes related to age of symptom onset with estimated age at %p-tau217 positivity and baseline age in progressors

Two outcomes were examined in the regression models: 1) time from baseline until symptom onset, which removes the birth-to-baseline age component; and 2) age at symptom onset, which shares the age scale with baseline age and estimated age at %p-tau217 positivity. Baseline age and estimated age at %p-tau217 positivity showed moderate correlations across all cohort/method combinations (Pearson r = 0.607–0.736, Spearman ρ = 0.502–0.642; **Supplementary Table 2**), indicating meaningful shared variance but sufficient independence to justify inclusion of both predictors in models.

The time from baseline until symptom onset was calculated by subtracting baseline age from the age at symptom onset. For this outcome, the second predictor was time from estimated %p-tau217 positivity to baseline, which represents years from %p-tau217 positivity at the time of the baseline visit. In the Knight ADRC/TIRA progressors, the associations with time from baseline until symptom onset for baseline age and time from %p-tau217 positivity to baseline were not significantly different (Adj. R^2^ 0.200 versus 0.285, p=0.546, **Figure 3A**). However, combining both predictors improved associations compared to baseline age alone (Adj. R^2^ 0.409 versus 0.200, p<0.001). Results were similar in the Knight ADRC/SILA progressors (**Supplementary Figure 5A**). In ADNI, associations between time from baseline until symptom onset and all predictors were weak, with all models yielding Adj. R^2^ below 0.18 (**Supplementary Figures 6A and 7A**).

**Figure 3.**
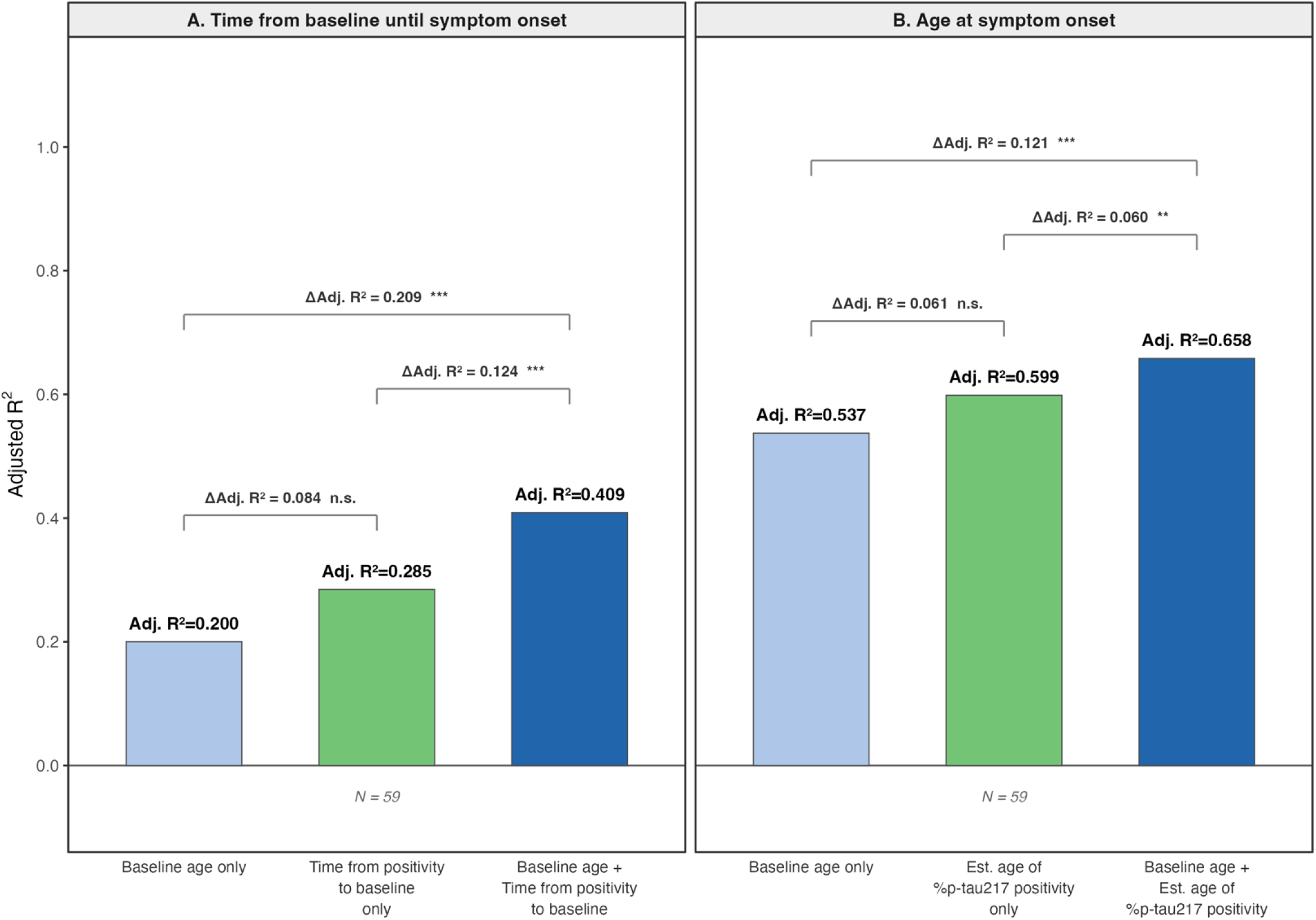
Correlations with outcomes related to symptom onset in progressors (Knight ADRC/TIRA). Time from baseline until symptom onset was correlated with baseline age, estimated time from %p-tau217 positivity to baseline, or the combination of both predictors **(A)**. Age at symptom onset was correlated with baseline age, estimated age at %p-tau217 positivity, or the combination of both predictors **(B)**. Asterisks indicate significance of the incremental contribution of the second predictor to the combined model: ***p<0.001, **p<0.01, *p<0.05; n.s., not significant.

In the Knight ADRC/TIRA progressors, baseline age explained much of the age at symptom onset (Adj. R^2^ 0.537) (**Figure 3B**). However, combining estimated age at %p-tau217 positivity with baseline age improved associations with age at symptom onset compared to baseline age alone, demonstrating that estimated age at %p-tau217 positivity has a unique contribution (Adj. R^2^ 0.658 versus 0.537, p<0.001). Results were similar in the Knight ADRC/SILA progressors, (**Supplementary Figure 5B**). With the ADNI/TIRA progressors, baseline age was more strongly associated with age at symptom onset than estimated age at %p-tau217 positivity by TIRA, and combining estimated age at %p-tau217 positivity with baseline age did not improve associations (**Supplementary Figure 6B**). However, with the ADNI/SILA progressors, combining estimated age at %p-tau217 positivity with baseline age improved associations with age at symptom onset compared to baseline age alone (Adj. R^2^ 0.678 versus 0.613, p=0.036; **Supplementary Figure 7B**).

To examine whether associations with age at symptom onset were stable or susceptible to design-related associations, we evaluated how the Adj. R^2^ changed as a function of follow-up restriction. The association of estimated age at %p-tau217 positivity with age at symptom onset was more stable as follow-up was truncated in both the Knight ADRC and ADNI cohorts, while the association of baseline age became more inflated as follow-up was truncated (**Supplementary Figure 8**).

### Variance in outcomes related to symptom onset associated with estimated age at %p-tau217 positivity and baseline age in progressors

Commonality analyses in the progressors partitioned the variance in each outcome related to symptom onset into components unique to estimated age at %p-tau217 positivity or time from %p-tau217 positivity to baseline, baseline age, and variance common to both predictors.

For time from baseline until symptom onset, time from %p-tau217 positivity to baseline contributed more unique variance than baseline age in the Knight ADRC/TIRA progressors (unique R^2^ 0.22 or 50.2% of total versus unique R^2^ 0.13 or 30.8% of total; **Table 1A**), with a smaller shared component (R^2^ 0.08 or 19.0% of total). The full model yielded an Adj. R^2^ of 0.41. Results were similar with the SILA model. In ADNI, associations were weak overall, with Adj. R^2^ of -0.09 for TIRA and 0.15 for SILA (**Table 1A**).

**Table 1.** Commonality analysis of associations between estimated age at %p-tau217 positivity and baseline age with outcomes related to symptom onset. R^2^ values reflect the proportion of total variance in each outcome attributable to each effect. % Total indicates each component as a percentage of the total R^2^. Adj. R^2^ is from the full two-predictor model.

**A. Time from baseline until symptom onset**
| Effect | Knight ADRC |  |  |  | ADNI |  |  |  |
| --- | --- | --- | --- | --- | --- | --- | --- | --- |
|  | TIRA, n=59 |  | SILA, n=61 |  | TIRA, n=20 |  | SILA, n=22 |  |
|  | R<sup>2</sup> | % Total | R<sup>2</sup> | % Total | R<sup>2</sup> | % Total | R<sup>2</sup> | % Total |
| Unique to time from %p-tau217 positivity to baseline | 0.22 | 50.2% | 0.22 | 47.1% | 0.02 | 97.3 | 0.21 | 90.9% |
| Unique to baseline age | 0.13 | 30.8% | 0.13 | 29.0% | 0.00 | 0.0 | 0.01 | 4.8% |
| Common to both | 0.08 | 19.0% | 0.11 | 23.9% | 0.00 | 2.7 | 0.01 | 4.4% |
| Total R<sup>2</sup> | 0.43 | 100.0% | 0.46 | 100.0% | 0.02 | 100.0 | 0.23 | 100.0% |
| Adj. R<sup>2</sup> | 0.41 |  | 0.44 |  | -0.09 |  | 0.15 |  |

**B. Age at symptom onset**
| Effect | Knight ADRC |  |  |  | ADNI |  |  |  |
| --- | --- | --- | --- | --- | --- | --- | --- | --- |
|  | TIRA, n=59 |  | SILA, n=61 |  | TIRA, n=20 |  | SILA, n=22 |  |
|  | R<sup>2</sup> | % Total | R<sup>2</sup> | % Total | R<sup>2</sup> | % Total | R<sup>2</sup> | % Total |
| Unique to estimated age of %p-tau217 positivity | 0.12 | 18.6% | 0.13 | 19.2% | 0.01 | 0.8% | 0.08 | 11.0% |
| Unique to baseline age | 0.06 | 9.6% | 0.06 | 8.2% | 0.36 | 49.5% | 0.21 | 30.2% |
| Common to both | 0.48 | 71.8% | 0.49 | 72.6% | 0.37 | 49.7% | 0.42 | 58.9% |
| Total R<sup>2</sup> | 0.67 | 100.0% | 0.67 | 100.0% | 0.74 | 100.0% | 0.71 | 100.0% |
| Adj. R<sup>2</sup> | 0.66 |  | 0.66 |  | 0.70 |  | 0.68 |  |

For age at symptom onset, estimated age at %p-tau217 positivity contributed more unique variance than baseline age in the Knight ADRC/TIRA progressors (unique R^2^ 0.12 or 18.6% of total versus unique R^2^ 0.06 or 9.6% of total; **Table 1B**), with the majority of variance shared between the two predictors (R^2^ 0.48 or 71.8% of total). The full model yielded an Adj. R^2^ of 0.66. Results were nearly identical with the SILA model. In ADNI, baseline age accounted for a larger proportion of unique variance than estimated age at %p-tau217 positivity by TIRA (unique R^2^ 0.36 or 49.5% of total versus unique R^2^ 0.01 or 0.8% of total) and SILA (unique R^2^ 0.21 or 30.2% versus unique R^2^ 0.08 or 11.0%; **Table 1B**).

### Randomization analyses of associations between outcomes related to symptom onset and estimated age at %p-tau217 positivity or baseline age in progressors

To examine whether observed associations could be explained by design alone, variables were randomly shuffled between individuals. The shuffling procedures varied by predictor-outcome combination to preserve relevant mathematical relationships. Both baseline age and either estimated age at %p-tau217 positivity or time from %p-tau217 positivity to baseline were tested as predictors.

For time from baseline until symptom onset, associations with time from %p-tau217 positivity to baseline substantially exceeded the null in the Knight ADRC for both TIRA (Adj R^2^ 0.285 versus 0.082, p<0.01) and SILA (Adj R^2^ 0.313 versus 0.101, p<0.01) (**Table 2A**). In ADNI, associations with time from %p-tau217 positivity to baseline did not exceed the null for TIRA (p=0.755) but were significant for SILA (Adj R^2^ 0.177 versus -0.008, p=0.017).

**Table 2.** Randomization analysis of outcomes related to symptom onset as a function of shuffled predictor values (10,000 permutations). For each predictor and outcome combination, shuffling procedures were tailored to preserve relevant mathematical relationships under the null. Observed adjusted R^2^ is from a simple linear regression of each outcome on the predictor in the unshuffled data. Permuted adjusted R^2^ is reported as mean (95% CI) of the null distribution with an empirical two-sided p-value, reflecting the proportion of permuted values whose absolute value equaled or exceeded the absolute observed adjusted R^2^. Analyses were performed separately in each dataset.

A. Time from baseline until symptom onset
|  | Knight ADRC<br>TIRA (n=59) |  |  | Knight ADRC<br>SILA (n=61) |  |  | ADNI<br>TIRA (n=20) |  |  | ADNI<br>SILA (n=22) |  |  |
| --- | --- | --- | --- | --- | --- | --- | --- | --- | --- | --- | --- | --- |
| Predictor | Obs.<br>Adj. $R^2$ | Null mean<br>(95% CI) | p-value | Obs.<br>Adj. $R^2$ | Null mean<br>(95% CI) | p-value | Obs.<br>Adj. $R^2$ | Null mean<br>(95% CI) | p-value | Obs.<br>Adj. $R^2$ | Null mean<br>(95% CI) | p-value |
| Time from<br>estimated %p-tau217 positivity<br>to baseline | 0.285 | 0.082<br>(-0.005–0.210) | <0.01 | 0.313 | 0.101<br>(0.007–0.233) | <0.01 | -0.032 | -0.012<br>(-0.056–0.159) | 0.755 | 0.177 | -0.008<br>(-0.050–0.153) | 0.017 |

B. Age at symptom onset
|  | Knight ADRC<br>TIRA (n=59) |  |  | Knight ADRC<br>SILA (n=61) |  |  | ADNI<br>TIRA (n=20) |  |  | ADNI<br>SILA (n=22) |  |  |
| --- | --- | --- | --- | --- | --- | --- | --- | --- | --- | --- | --- | --- |
| Predictor | Obs.<br>Adj. $R^2$ | Null mean<br>(95% CI) | p-value | Obs.<br>Adj. $R^2$ | Null mean<br>(95% CI) | p-value | Obs.<br>Adj. $R^2$ | Null mean<br>(95% CI) | p-value | Obs.<br>Adj. $R^2$ | Null mean<br>(95% CI) | p-value |
| Estimated age of<br>%p-tau217<br>positivity (direct<br>shuffle) | 0.599 | -0.000<br>(-0.018–0.067) | <0.001 | 0.612 | 0.000<br>(-0.017–0.066) | <0.001 | 0.337 | 0.000<br>(-0.056–0.209) | <0.01 | 0.470 | 0.000<br>(-0.050–0.185) | <0.001 |
| Estimated age of<br>%p-tau217<br>positivity (timing<br>shuffled) | 0.599 | 0.319<br>(0.187–0.464) | <0.001 | 0.612 | 0.333<br>(0.205–0.471) | <0.001 | 0.337 | 0.210<br>(-0.030–0.530) | 0.418 | 0.470 | 0.239<br>(0.006–0.534) | 0.084 |
| Baseline age<br>(direct shuffle) | 0.537 | 0.000<br>(-0.018–0.068) | <0.001 | 0.537 | 0.000<br>(-0.017–0.068) | <0.001 | 0.714 | 0.000<br>(-0.055–0.213) | <0.001 | 0.613 | 0.001<br>(-0.050–0.192) | <0.001 |

For age at symptom onset, associations with estimated age at %p-tau217 positivity exceeded the null across all four datasets when positivity age was shuffled directly (all p<0.01; **Table 2B**). When the null was instead constructed by shuffling timing while preserving the baseline age anchor, estimated age at %p-tau217 positivity still substantially exceeded the null in the Knight ADRC for both TIRA (Adj R^2^ 0.599 versus 0.319, p<0.001) and SILA (Adj R^2^ 0.612 versus 0.333, p<0.001), although results were not significant in ADNI (TIRA p=0.418; SILA p=0.084). Associations with baseline age exceeded the null in all four datasets (p<0.001).

## Discussion

This study used complementary analytical approaches to examine whether estimated age at plasma %p-tau217 positivity, derived from plasma %p-tau217 clock models, is less affected than baseline age by design-related associations when evaluating the risk of progression and timing of symptomatic Alzheimer’s disease (AD). In analyses including both progressors and non-progressors from the Knight ADRC and ADNI, estimated age at %p-tau217 positivity demonstrated superior discrimination for ranking age of onset of AD symptoms. In progressor sub-cohorts, regression and commonality analyses overall showed that estimated age at %p-tau217 positivity contributed unique variance beyond baseline age in explaining the timing of symptom onset. Additionally, randomization analyses found that associations between estimated %p-tau217 positivity and symptom onset exceeded those expected by chance in the larger Knight ADRC cohort, with some mixed results in the smaller ADNI cohort.

Several lines of evidence support the complementary and added value of the clock-derived estimate to baseline age in studies of clinical progression with limited follow-up. The clock approach, by anchoring to a distinct biological event (%p-tau217 positivity) that often precedes the baseline plasma collection, may enable analyses of a greater number of clinical progressors in research studies when plasma data are not available from the baseline visit. Further, the clock approach may extend the effective observation window, which may mitigate design-associated inflation of associations that occurs with short follow-up. The clock approach yields longer median intervals to symptom onset (approximately 6.5–12.4 years versus 2.4–6.0 years from baseline). In C-index analyses including all initially cognitively unimpaired participants, estimated age at %p-tau217 positivity consistently outperformed baseline age in ordering progression to an AD clinical syndrome or to symptomatic AD when all available follow-up was considered. As clinical follow-up was progressively truncated, the C-index for estimated age at %p-tau217 positivity remained stable across all observation windows, while the C-index for baseline age increased, particularly at the shortest observation windows (≤5 and ≤2 years). At shorter observation windows, only participants who converted quickly remain as events, concentrating the comparable pairs among a selected subset in which baseline age and symptom onset age are more tightly linked by the study design. This pattern was more pronounced in ADNI, where the C-index for baseline age approached that of estimated age at %p-tau217 positivity at the shortest windows.

In the progressor sub-cohorts, estimated age at %p-tau217 was additive with baseline age in associations with symptom onset outcomes. For time from baseline until symptom onset, associations were weaker but the time from estimated %p-tau217 positivity to baseline contributed more unique variance than baseline age. When predicting age at symptom onset, baseline age and estimated age at %p-tau217 positivity shared a large proportion of explained variance, reflecting overlap in the age scale. However, the estimated age at %p-tau217 positivity provided incremental predictive value beyond baseline age in the larger Knight ADRC cohort. Mixed results in the smaller ADNI cohorts may reflect lower statistical power due to the smaller numbers of progressors and shorter follow-up, resulting in inflation of the effect of baseline age. While these results might suggest that models predicting symptom onset would improve if both estimated age at %p-tau217 positivity and baseline age were included, truncation analyses suggest that baseline age is more susceptible to design-related associations and therefore the accuracy of models including baseline age could decrease with shorter follow-up.

Permutation-based randomization analyses tested whether associations exceeded what would be expected from a structurally similar but uninformative predictor. For time from baseline until symptom onset, associations with estimated %p-tau217 positivity exceeded the null in the Knight ADRC but results were mixed in ADNI. For age at symptom onset, associations with age at %p-tau217 positivity exceeded the null across all four datasets when age at %p-tau217 positivity was shuffled directly. If age at %p-tau217 positivity represents a distinct biological event that is not directly coupled with baseline age, as is suggested by results showing that age at %p-tau217 positivity is highly correlated with observed age at conversion to %p-tau217 positivity ^7^ and differential changes between age at %p-tau217 positivity and baseline age with follow-up truncation, this null is more appropriate. If age at %p-tau217 positivity is considered mathematically coupled to baseline age, a more conservative null that preserves the baseline age anchor of the predictor is more appropriate. With the more conservative null, associations between estimated age at %p-tau217 positivity and age at symptom onset were significantly higher in the Knight ADRC, but were not significant in ADNI. Associations between baseline age and age at symptom onset exceeded the null across all datasets. Overall, these results support that estimated %p-tau217 positivity captures biological information about the timing of symptom onset beyond design-related associations.

This study had multiple limitations and considerations for future directions. Most importantly, as was clearly stated and discussed in the Petersen *et al*. paper, these models are currently only relevant to group-level research studies and clinical trials, not individual-level use ^7^. Analyses of the timing of symptom onset in progressor sub-cohorts are, by definition, conditional on observed progression during follow-up and cannot directly evaluate absolute error or calibration in non-progressors. Models that use continuous rather than categorical measures of cognition may enable both progressors and non-progressors to be considered ^10^. The number of progressors was low, particularly in ADNI (n=20/22), which limits statistical power and generalizability and may explain the differences in findings between Knight ADRC and ADNI. Prospective validation in independent cohorts is needed to evaluate whether models of symptom onset created in progressors are generalizable. Further, integrating biomarker clocks into trial enrichment strategies will require careful validation and calibration against clinical endpoints. Another limitation is the error of estimated age at %p-tau217 from the clock models, which may affect the associations with outcomes related to progression. Development of methods that propagate uncertainty related to the %p-tau217 assay and modeling approach and competing-risk frameworks that account for death would strengthen inferences. Multi-biomarker clocks that include measures of co-pathologies and approaches that integrate longitudinal biomarker trajectories may also improve predictions ^10^. Sharing of code, data, and web-based applications will accelerate independent replication and methodological refinement across research groups. Additional limitations include that both cohorts were predominantly non-Hispanic White and highly educated. Future studies should examine larger, more diverse longitudinal cohorts with extended clinical follow-up and higher progression rates.

In conclusion, estimated age at %p-tau217 positivity derived from plasma clock models offers a biologically anchored metric that may be less susceptible to design-induced associations than baseline age and capture unique information about the timing of symptomatic AD. These properties are most evident in the larger Knight ADRC cohort. While challenges remain in modeling the transition from preclinical to symptomatic AD, these findings support the utility of plasma p-tau217 clock models as a valuable complement to conventional baseline-anchored analyses in preclinical AD research and trial design.

## Data Availability

Data from the Knight ADRC can be requested by qualified investigators (https://knightadrc.wustl.edu/professionals-clinicians/request-center-resources/). Data from ADNI can be requested via the LONI website (adni.loni.usc.edu). All analyses were performed in R (version 4.4.1). Codes used in these analyses are available from https://github.com/WashUFluidBiomarkers/design-related-associations-in-AD-clock-models.

https://adni.loni.usc.edu/

https://github.com/WashUFluidBiomarkers/design-related-associations-in-AD-clock-models

## Acknowledgments

We thank Phillip Insel, Michael Donohue, Susan Landau, and William Jagust for their constructive comments. These analyses used, in part, code shared by Philip Insel and Michael Donohue. Funding for this data re-analysis was supported by National Institute on Aging grant R01AG070941 (SES).

## References

1. Yaari R, Holdridge KC, Williamson M, et al. Donanemab in preclinical Alzheimer’s disease: Screening and baseline data from TRAILBLAZER-ALZ 3. Alzheimer’s & dementia : the journal of the Alzheimer’s Association 2025;21:e70662.

2. Rafii MS, Sperling RA, Donohue MC, et al. The AHEAD 3-45 Study: Design of a prevention trial for Alzheimer’s disease. Alzheimer’s & dementia : the journal of the Alzheimer’s Association 2023;19:1227–1233.

3. McDade E, Llibre-Guerra JJ, Holtzman DM, Morris JC, Bateman RJ. The informed road map to prevention of Alzheimer Disease: A call to arms. Mol Neurodegener 2021;16:49.

4. Bateman RJ, Benzinger TL, Berry S, et al. The DIAN-TU Next Generation Alzheimer’s prevention trial: Adaptive design and disease progression model. Alzheimer’s & dementia : the journal of the Alzheimer’s Association 2017;13:8–19.

5. Ryman DC, Acosta-Baena N, Aisen PS, et al. Symptom onset in autosomal dominant Alzheimer disease: a systematic review and meta-analysis. Neurology 2014;83:253–260.

6. Li Y, Yen D, Hendrix RD, et al. Timing of Biomarker Changes in Sporadic Alzheimer’s Disease in Estimated Years from Symptom Onset. Annals of neurology 2024.

7. Petersen KK, Mila-Aloma M, Li Y, et al. Predicting onset of symptomatic Alzheimer’s disease with plasma p-tau217 clocks. Nat Med 2026;32:1085–1094.

8. Insel PS, Donohue MC. Design-induced artifacts when “disease clocks” are plugged into second-stage analyses of symptom onset. medRxiv 2026:2026.2003.2026.26349230.

9. Winkler AM, Ridgway GR, Webster MA, Smith SM, Nichols TE. Permutation inference for the general linear model. NeuroImage 2014;92:381–397.

10. Ohman F, Raket LL, Scholl M, Alzheimer’s Disease Neuroimaging I. Cognitive Trajectories from Preclinical Alzheimer’s Disease to Dementia. Adv Sci (Weinh) 2026:e18124.

