## Supplementary information for "Time until symptoms and design-related associations in Alzheimer’s disease clinical progression analyses"

Kellen K. Petersen, Yan Li, Suzanne E. Schindler

**Supplementary information**

Supplementary Table 1. Paired comparison of time from baseline age to symptom onset versus time from estimated age at %p-tau217 positivity to symptom onset.

Supplementary Table 2. Pearson and Spearman correlations between baseline age and estimated age at %p-tau217 positivity in progressor subsets.

Supplementary Figure 1. Concordance of age at symptom onset with age at %p-tau217 positivity or baseline age (Knight ADRC/SILA).

Supplementary Figure 2. Concordance of age at symptom onset with age at %p-tau217 positivity or baseline age (ADNI/TIRA).

Supplementary Figure 3. Concordance of age at symptom onset with age at %p-tau217 positivity or baseline age (ADNI/SILA).

Supplementary Figure 4. Concordance of age at symptom onset with age at %p-tau217 positivity or baseline age (Knight ADRC/TIRA).

Supplementary Figure 5. Correlations with outcomes related to symptom onset in progressors (Knight ADRC/SILA).

Supplementary Figure 6. Correlations with outcomes related to symptom onset in progressors (ADNI/TIRA).

Supplementary Figure 7. Correlations with outcomes related to symptom onset in progressors (ADNI/SILA).

Supplementary Figure 8. Sensitivity of associations between age at symptom onset and predictors to follow-up restriction.

**Supplementary Table 1. Paired comparison of time from baseline age to symptom onset versus time from estimated age at %p-tau217 positivity to symptom onset.** Within each cohort/method combination, a one-sided paired Wilcoxon signed-rank test was used to evaluate whether time from estimated age at %p-tau217 positivity to symptom onset was significantly longer than time from baseline age to symptom onset. The median difference is positivity-to-onset minus baseline-to-onset. p-values reflect the one-sided alternative that positivity-to-onset is greater.

| **Cohort** | **Method** | **Symptoms before baseline,**  **n (%)** | **Median time from baseline to onset,**  **years (IQR)** | **Median time from positivity to onset, years (IQR)** | **Median difference,  years (IQR)** | **p-value** |
| --- | --- | --- | --- | --- | --- | --- |
| **Knight ADRC** | **TIRA** | 8 (13.6%) | 5.8 (1.1-10.0) | 9.4 (6.2-13.5) | 5.0 (0.7-8.8) | <0.0001 |
| **Knight ADRC** | **SILA** | 7 (11.5%) | 6.0 (1.1-10.0) | 9.1 (5.1-12.3) | 3.5 (-0.5-7.2) | 0.0001 |
| **ADNI** | **TIRA** | 3 (15.0%) | 2.4 (1.4-5.0) | 12.4 (7.4-15.9) | 10.7 (6.2-14.4) | 0.0002 |
| **ADNI** | **SILA** | 3 (13.6%) | 3.0 (1.7-5.5) | 6.5 (4.1-9.6) | 5.8 (0-7.8) | 0.0039 |

**Supplementary Table 2. Pearson and Spearman correlations between baseline age and estimated age at %p-tau217 positivity in progressor subsets.** Pearson r and Spearman ρ were computed between baseline age (age at baseline plasma sample) and estimated age at %p-tau217 positivity for each cohort/method combination. Analyses were conducted in progressor subsets from the Knight ADRC and ADNI. Spearman p-values were computed using exact permutation methods.

| **Cohort** | **Method** | **Pearson** | | **Spearman** | |
| --- | --- | --- | --- | --- | --- |
|  |  | **r** | **p** | **ρ** | **p** |
| Knight ADRC | TIRA | 0.724 | <0.0001 | 0.642 | <0.0001 |
| Knight ADRC | SILA | 0.736 | <0.0001 | 0.609 | <0.0001 |
| ADNI | TIRA | 0.645 | 0.0021 | 0.571 | 0.0096 |
| ADNI | SILA | 0.607 | 0.0028 | 0.502 | 0.0185 |

**Supplementary Figure 1. Concordance of age at symptom onset with age at %p-tau217 positivity or baseline age (Knight ADRC/SILA).** For individuals who were cognitively unimpaired at their baseline cognitive assessment, including both progressors and non-progressors, the ability of baseline age and estimated age at %p-tau217 positivity to rank age of onset of either (**A**) symptomatic AD (an AD syndrome with estimated %p-tau217 positivity) or (B) an AD syndrome were evaluated. Follow-up was truncated at fixed intervals from the baseline plasma sample to simulate trial observation windows of varying duration. The C-index was estimated using interval-censoring with 2000-sample bootstrapped 95% confidence intervals. Sample sizes and event counts at each restriction level are shown below the x-axis.

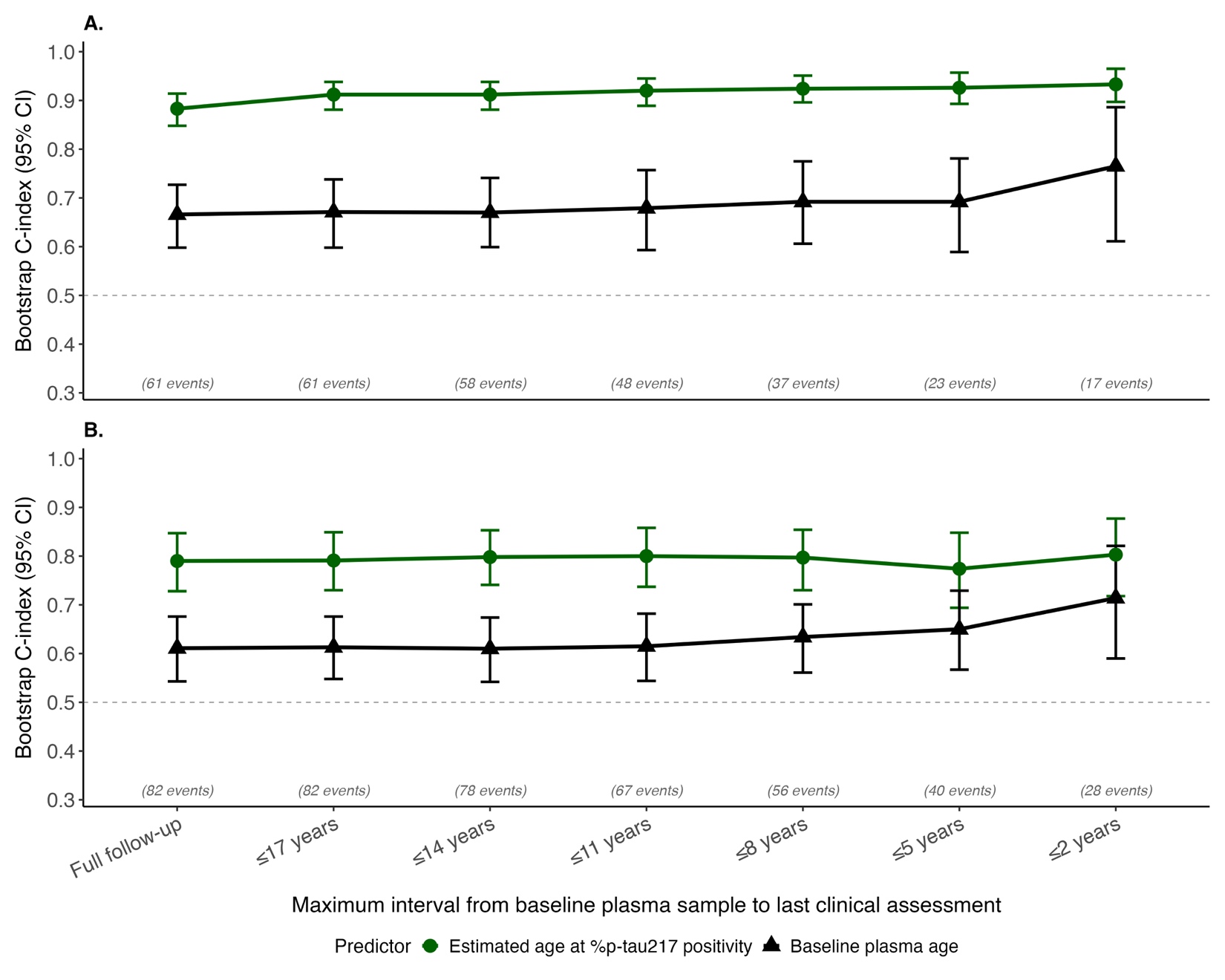

**Supplementary Figure 2. Concordance of age at symptom onset with age at %p-tau217 positivity or baseline age (ADNI/TIRA).** For individuals who were cognitively unimpaired at their baseline cognitive assessment, including both progressors and non-progressors, the ability of baseline age and estimated age at %p-tau217 positivity to rank age of onset of either (**A**) symptomatic AD (an AD syndrome with estimated %p-tau217 positivity) or (B) an AD syndrome were evaluated. Follow-up was truncated at fixed intervals from the baseline plasma sample to simulate trial observation windows of varying duration. The C-index was estimated using interval-censoring with 2000-sample bootstrapped 95% confidence intervals. Sample sizes and event counts at each restriction level are shown below the x-axis.

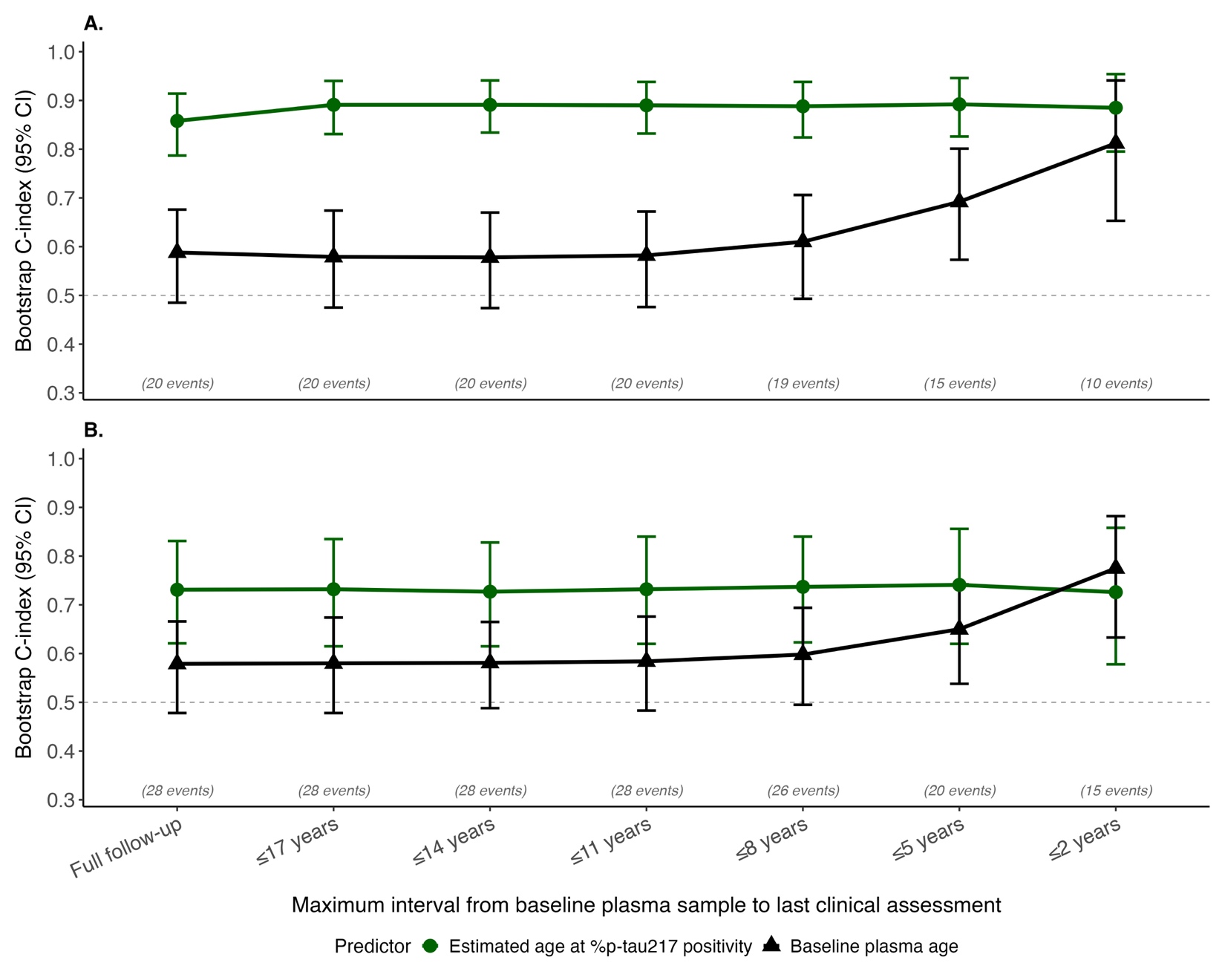

**Supplementary Figure 3. Concordance of age at symptom onset with age at %p-tau217 positivity or baseline age (ADNI/SILA).** For individuals who were cognitively unimpaired at their baseline cognitive assessment, including both progressors and non-progressors, the ability of baseline age and estimated age at %p-tau217 positivity to rank age of onset of either (**A**) symptomatic AD (an AD syndrome with estimated %p-tau217 positivity) or (B) an AD syndrome were evaluated. Follow-up was truncated at fixed intervals from the baseline plasma sample to simulate trial observation windows of varying duration. The C-index was estimated using interval-censoring with 2000-sample bootstrapped 95% confidence intervals. Sample sizes and event counts at each restriction level are shown below the x-axis.

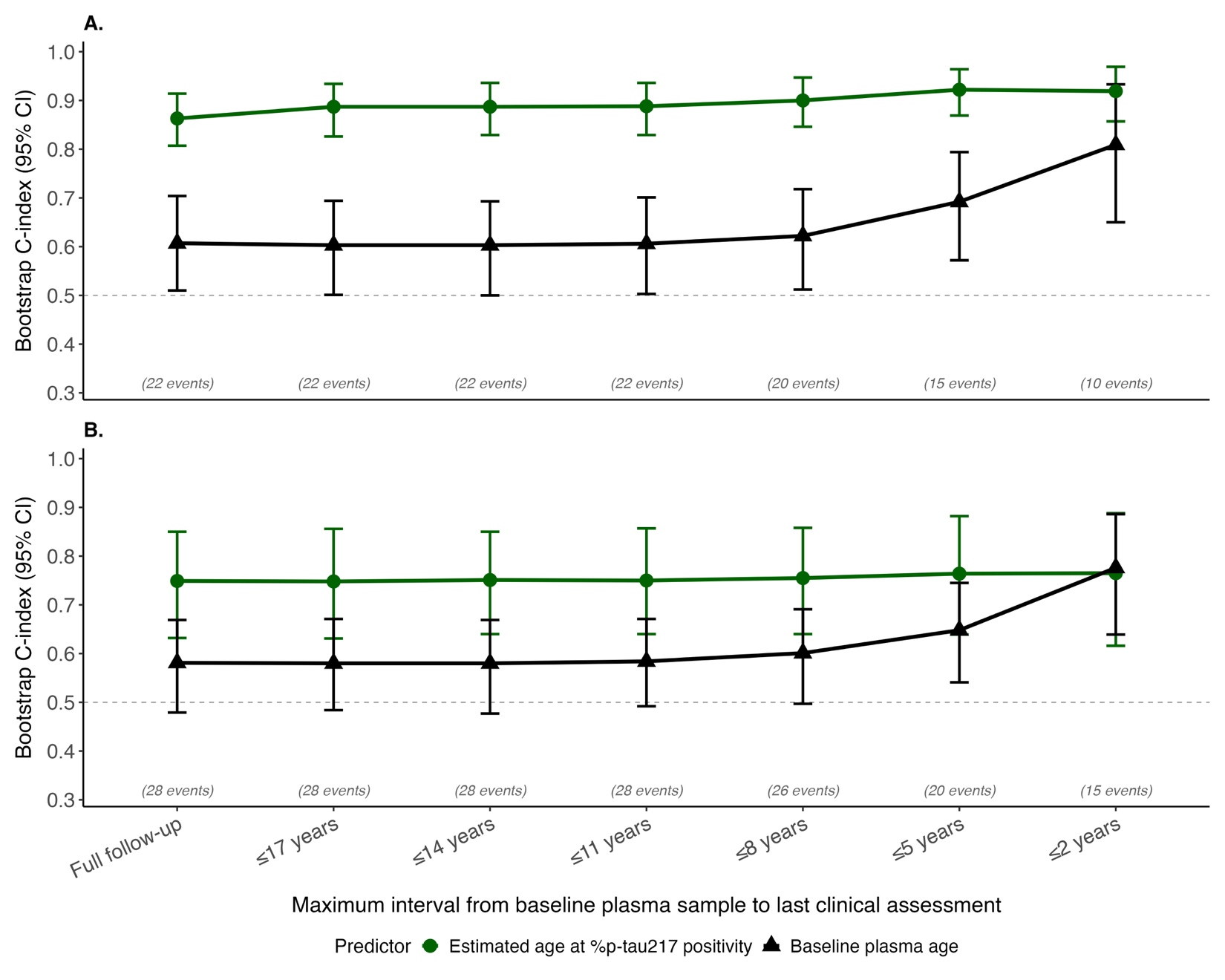

**Supplementary Figure 4. Concordance of age at symptom onset with age at %p-tau217 positivity or baseline age (Knight ADRC/TIRA).** For individuals who were cognitively unimpaired at their baseline cognitive assessment, including both progressors and non-progressors, the ability of baseline age and estimated age at %p-tau217 positivity to rank age of onset of either (**A**) symptomatic AD (an AD syndrome with estimated %p-tau217 positivity) or (B) an AD syndrome were evaluated. Follow-up from the baseline plasma sample was truncated by right-censoring the age of onset at the last assessment within the truncation interval for non-progressors or those who progressed after the truncation interval. The C-index was estimated using right-censoring with 2000-sample bootstrapped 95% confidence intervals. Sample sizes and event counts at each restriction level are shown below the x-axis. Similar results were obtained with the Knight ADRC/SILA, ADNI/TIRA, and ADNI/SILA datasets.

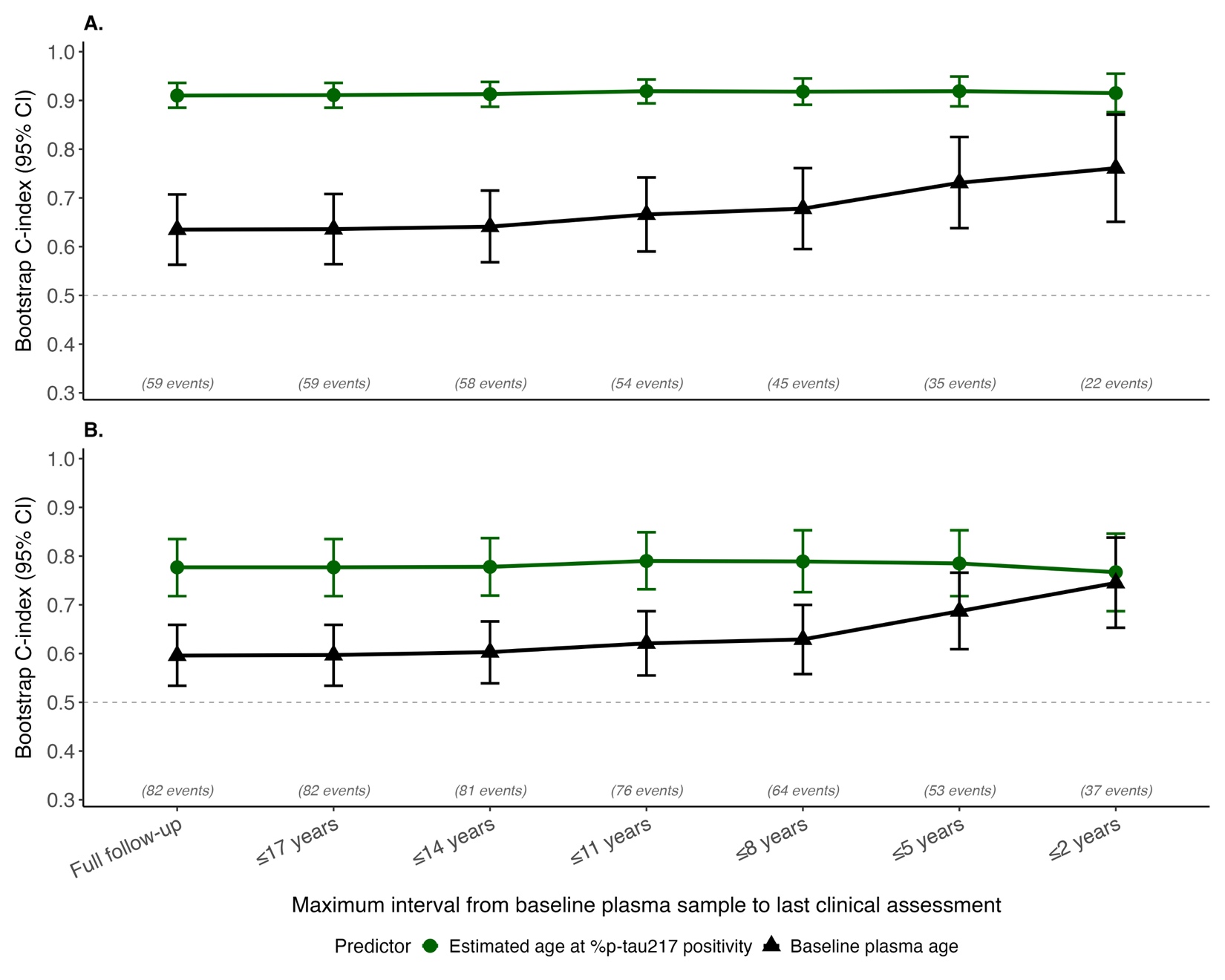

**Supplementary Figure 5. Correlations with outcomes related to symptom onset in progressors (Knight ADRC/SILA).** Time from baseline until symptom onset was correlated with baseline age, estimated time from %p-tau217 positivity to baseline, or the combination of both predictors (**A**). Age at symptom onset was correlated with baseline age, estimated age at %p-tau217 positivity, or the combination of both predictors (**B**). Asterisks indicate significance of the incremental contribution of the second predictor to the combined model: ***p<0.001, **p<0.01, *p<0.05; n.s., not significant.

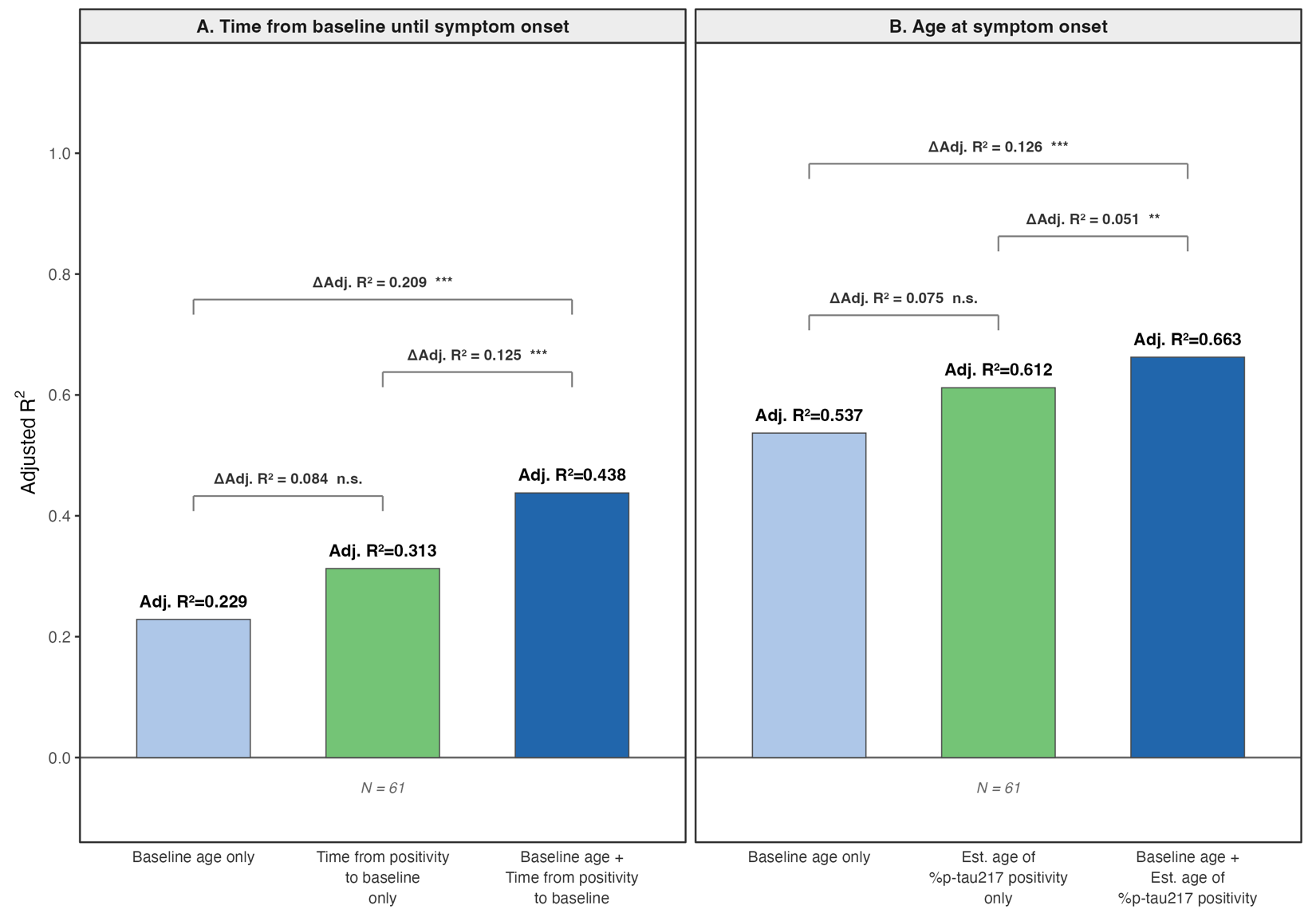

**Supplementary Figure 6. Correlations with outcomes related to symptom onset in progressors (ADNI/TIRA).** Time from baseline until symptom onset was correlated with baseline age, estimated time from %p-tau217 positivity to baseline, or the combination of both predictors (**A**). Age at symptom onset was correlated with baseline age, estimated age at %p-tau217 positivity, or the combination of both predictors (**B**). Asterisks indicate significance of the incremental contribution of the second predictor to the combined model: ***p<0.001, **p<0.01, *p<0.05; n.s., not significant.

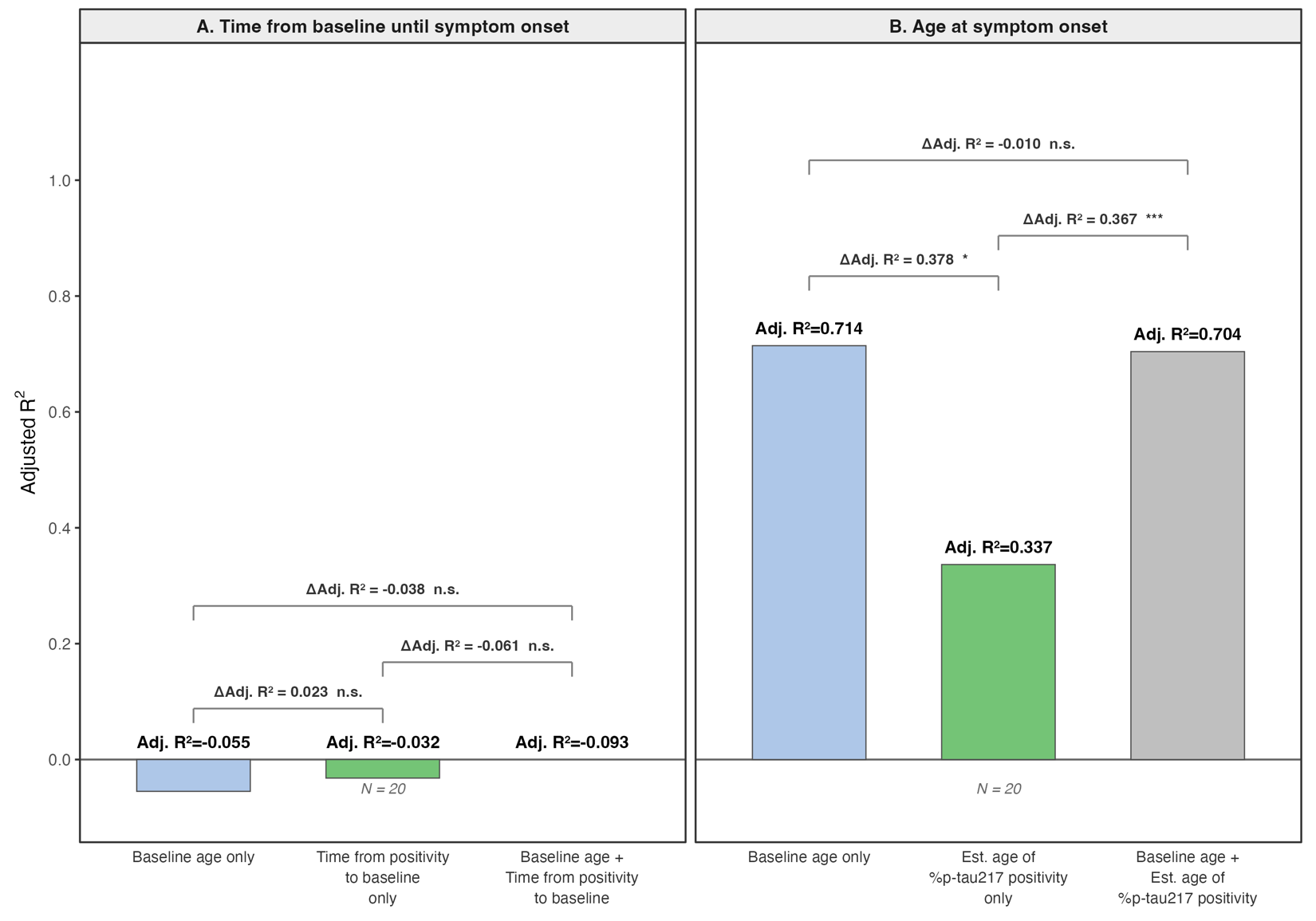

**Supplementary Figure 7. Correlations with outcomes related to symptom onset in progressors (ADNI/SILA).** Time from baseline until symptom onset was correlated with baseline age, estimated time from %p-tau217 positivity to baseline, or the combination of both predictors (**A**). Age at symptom onset was correlated with baseline age, estimated age at %p-tau217 positivity, or the combination of both predictors (**B**). Asterisks indicate significance of the incremental contribution of the second predictor to the combined model: ***p<0.001, **p<0.01, *p<0.05; n.s., not significant.

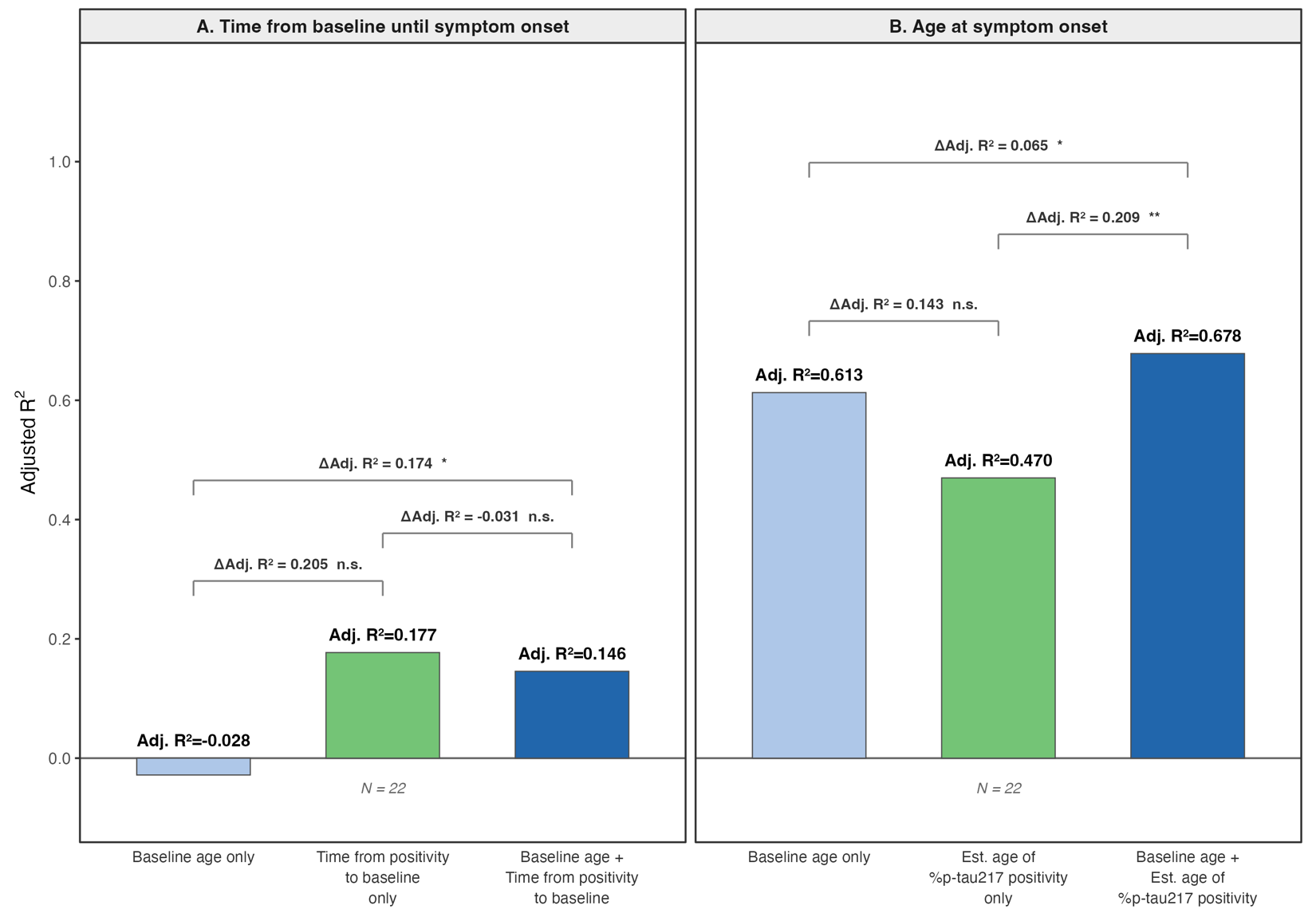

**Supplementary Figure 8. Sensitivity of associations between age at symptom onset and predictors to follow-up restriction.** The left panels show the adjusted R² from a simple linear regression of age at symptom onset as a function of estimated age at %p-tau217 positivity (green) and baseline plasma age (black) with progressively restricted time intervals from baseline to symptom onset. The right panels show the number of progressors included at each restriction level. Results are shown for progressors from the Knight ADRC/TIRA, Knight ADRC/SILA, ADNI/TIRA, and ADNI/SILA.

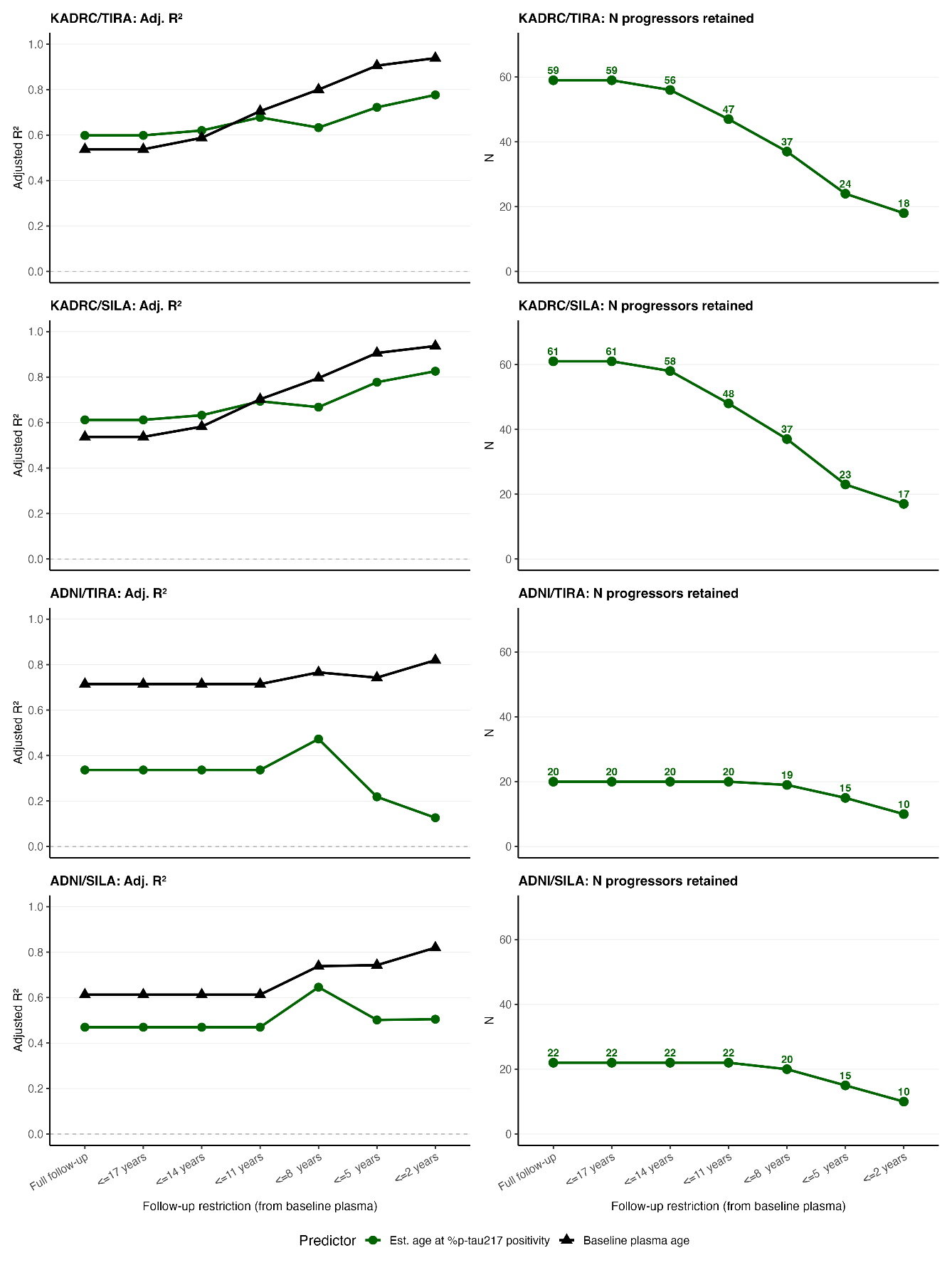
